# Performance of the *SD Bioline* rapid diagnostic test as a good alternative to the detection of Human African Trypanosomiasis in Cameroon

**DOI:** 10.1101/2022.05.10.22274663

**Authors:** Andrillene Laure Deutou Wondeu, Aline Okoko, Ghyslaine Bruna Djeunang Dongho, Christian Doll, Samuel Bahebegue, Ulrich Stéphane Mpeli Mpeli, Christian Chouamou Ninko, Carla Montesano, Nicolas Félicien Dologuele, Herman Parfait Awono Ambene

## Abstract

**Background:** The management of human African trypanosomiasis (HAT), caused by *Trypanosoma brucei gambiense* relies on case detection. As part of this, the routine screening by the card agglutination test for trypanosomiasis (CATT) is the critical step before parasitological confirmation. Individual rapid diagnostic tests (RDTs) have recently been developed for the serodiagnosis of HAT.

**Objective:** The objective of the current study was to assess the input of SD Bioline HAT on the serological screening of human African trypanosomiasis in Cameroonian foci.

**Methods:** Blood samples were collected during the surveys in the foci of Campo, Yokadouma and Fontem between June 2014 and January 2015. Diagnostic performance indicators such as sensitivity (Se) and specificity (Sp) of SD Bioline HAT was determined from the CATT, used as gold standard for the detection of specific antibodies of *Trypanosoma brucei gambiense*.

**Results:** A total of 88 samples were tested in Campo 59.1% (n=52), Yokadouma 31.8% (n=28) and Fontem 9.1% (n=8). The gender distribution was 61.4% (n=54) male and 38.4% (n=34) female, with an average age of 35.4 ± 19.0 years. The overall seroprevalence was 11.4% (95% CI: 6.3- 19.7) with the CATT method and 18.2% (95% CI: 11.5- 27.5) with the SD Bioline HAT RDT method in probed foci. The Se and Sp were 80.0% and 89.7% respectively for the SD Bioline HAT.

**Conclusion:** This study showed that the overall performance of the SD Bioline HAT was close to that of the CATT, with significant specificity in the serological detection of HAT.

## Introduction

Human African trypanosomiasis (HAT), also known as sleeping sickness, is a vector-borne parasitosis that is endemic in many countries of sub-Saharan Africa^1^. It is caused by a flagellated protozoan of the genus Trypanosoma, naturally transmitted to man by the Tsetse fly. HAT is one of the neglected and deadly tropical diseases with its Rhodesian form due to *Trypanosoma brucei rhodesiense* widespread in eastern Africa and the Gambian form caused by *Trypanosoma brucei gambiense* in central and western Africa ^2,3^. It is characterized by a non-specific clinical presentation with no consistent, pathognomonic manifestations ^4^.

HAT remains a worrying problem in intertropical Africa where it is currently on the upsurge ^5^. The World Health Organization (WHO) reported 3,797 cases of HAT in 2014 and 2,804 new cases in 2015. The disease caused an estimated 3,500 deaths in 2015.Sustained control efforts have reduced the number of new cases, as in 2019, there were 992 and 663 cases reported in 2020 cases recorded respectively and more than 95% of the reported cases were due to *T. b. gambiense* ^6,7^. In Central Africa, there were several outbreaks of endemicity, Democratic Republic of the Congo (DRC) is the most affected country, with more than 75% of the gambiense cases declared ^8^.

The management of the Gambian form is based on the detection of cases, followed by appropriate treatment according to the stage of the disease ^9^. The routine technique for mass screening is the CATT ^10^, however, the constraints of its use limit its application under certain conditions during active and passive screening ^11^. As a result, new strategies at the stages of fight and control of HAT have been established.

In recent years, rapid diagnostic tests (RDTs) have been developed and evaluated in some countries for the detection of HAT ^12,13^. These tests are an alternative choice for routine screening at the health facilities of endemic foci with the goal of HAT elimination by 2030 set by the WHO. Further studies on the performance of these RDTs are necessary for their effective dissemination. Therefore, we evaluated the performance of the SD Bioline HAT, the RDTs for the detection of HAT cases at *T.b. gambiense* in three foci (Campo, Fontem and Yokadouma) of HAT in Cameroon, where the National Program for Control of Human African Trypanosomiasis (PNLTHA) is actively investigating cases.

## Materials and Methods

### Ethical considerations

All necessary precautions were taken to ensure that the rights and freedoms of the participants in the research were respected. In order to conduct the present study, the ethical clearance N° 2015 / 0003 was sought and obtained from the institutional ethics committee for research for human health of the school of health sciences (Yaounde).

### Study design

We conducted a cross-sectional study over a period of nine months, from June 2014 to March 2015. The samples were collected in three foci: Campo in the South, Yokadouma in the East, and Fontem in the Southwest of Cameroon. These foci are geographically propitious zones for hatching Tsetse flies: a temperature of about 25°C and a relative humidity of 80 to 85% and a lot of shade ^14^. The participants in the study were the inhabitants of the foci and the refugees from the Central African Republic residing in the Yokadouma camp. However, pregnant women and infants were not included in our study.

Sampling was done by simple random sampling. It consisted in drawing lots directly from individuals in the population of the various foci surveyed.

### Participant enrolment process

Our work was conducted during the various surveys organized by the national HAT control program of Cameroon. They were done in several stages, ranging from the census to prior awareness of the target population by the field team. We then approached each participant by presenting the information notice, explaining to him in simple terms the purpose of the study, the interest, the amount of blood to be collected and the samples and results management.

Anyone who understood and accepted the conditions of the study gave their consent by signing the informed consent form. After this stage, each participant was registered and then taken to perform the different CATT and rapid diagnostic screening tests. At the end, the results were handed to them individually.

Blood sampling consisted in taking from each participant, on the fingertip, about 200 μl of blood. This blood was stored in heparin microcapillary tube and classified on racks numbered from 1 to 10 for the first step of screening. However, for all individuals who tested positive in the first step of screening, whole blood was collected from the elbow in a 4ml heparinised tube for further testing (CATT dilution and RDTs).

### Tests performed

The samples were analyzed both in the field and at the HAT laboratory at the Organisation for Coordination of the Control of Endemic Diseases in Central Africa, Yaounde. Two types of tests were performed for each sample: the agglutination test for trypanosomiasis and the rapid diagnostic test.

#### Card Agglutination Test for Trypanosomiasis: CATT (Figure 1)

The Card Agglutination Test for Trypanosomiasis is a direct plate agglutination test. It consists in bringing together antigens of trypanosomes, consisting of whole and lyophilized *T. b. gambiense*; and the whole blood of the person to be examined. It is an interaction between a specific agglutinating antibody and a particular antigen.

**Figure 1.**
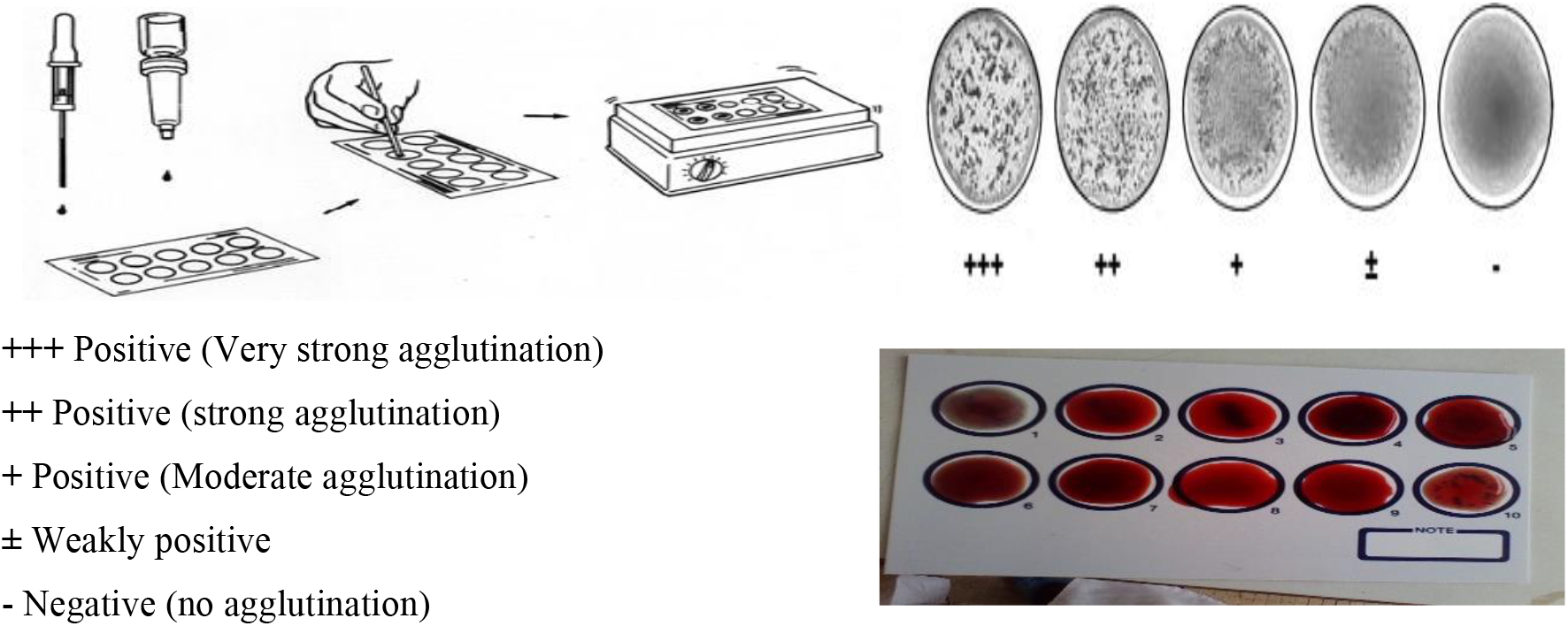
Stages of the CATT. (Test Guide CATT/*T.b.gambiense* Institute of Tropical Medicine) CATT: Card Agglutination Test for Trypanosomiasis

A drop of reconstituted reagent (about 45 μl) was deposited on the card, a drop of blood (about 25 μl) was added. The mixture was then spread, the card was placed five minutes on a rotary shaker at one revolution per second. The reading was done immediately after the 5 minutes of stirring, with the naked eye with reference to the positive and negative controls.

Quantitation was performed on all positive CATT whole blood samples. It consisted in taking 100 μl of whole blood for successive dilutions (1:2, 1:4, 1: 8, 1:16) with 100 μl CATT buffer each time. The titration was done by taking 25 μl of each blood dilution that we put in the test area on the card, before adding a drop of reconstituted reagent. For the rest we proceeded as for the CATT test. This quantification was done in order to determine the positivity threshold of each sample.

#### The rapid diagnostic test

The rapid diagnostic test SD Bioline HAT is an immunochromatographic test for the rapid and qualitative detection of antibodies named Litat1.3 and Litat1.5, which are specific for *T. b. gambiense*. The procedure used was according to the manufacturer’s instructions (Figure 2). A drop of whole blood was placed in the round window of the test. Subsequently, we added four (04) drops of diluent to this spot, the result was read within 15 minutes of the deposit.

**Figure 2.**
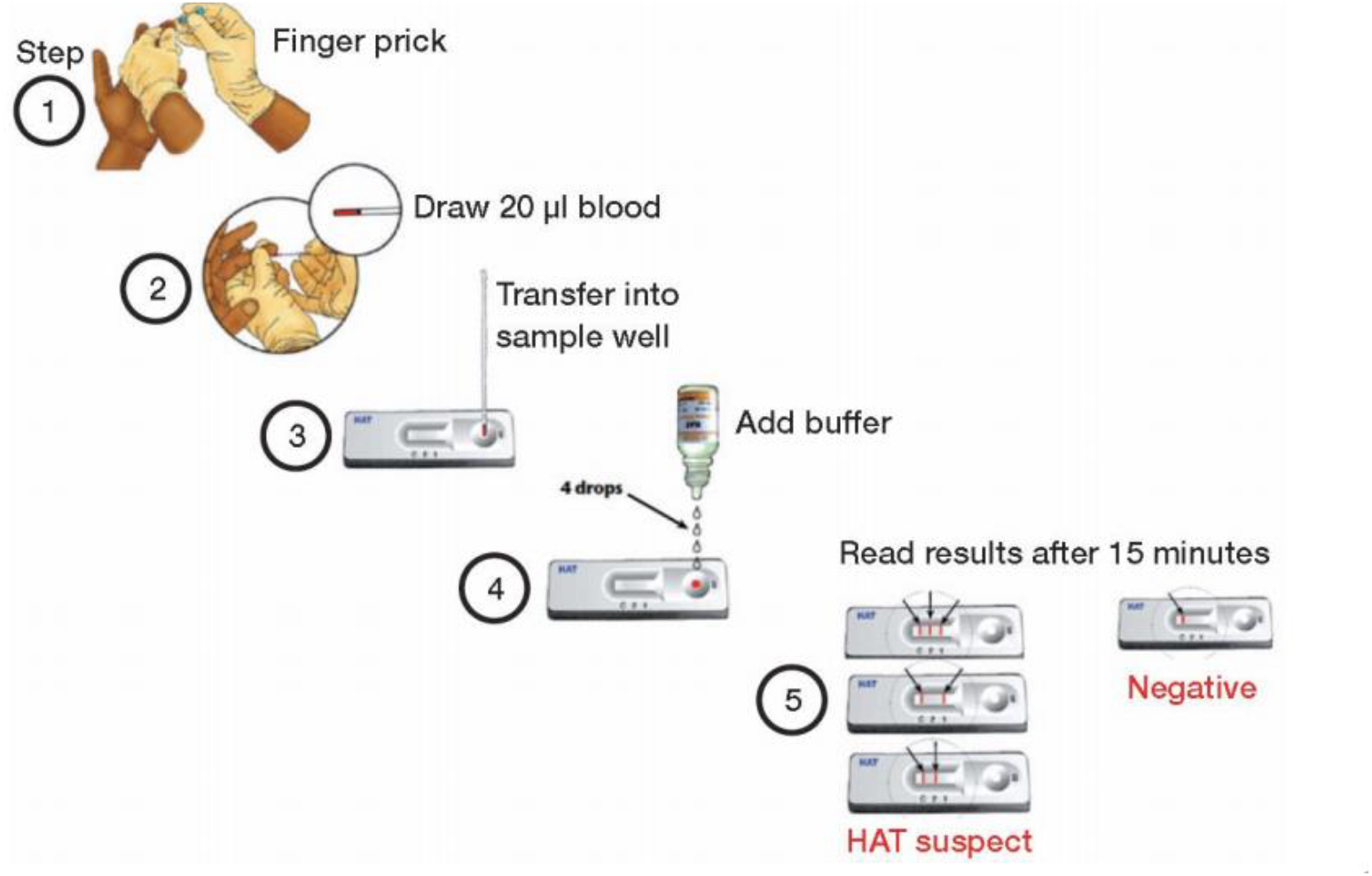
Procedure of the DS Bioline HAT test (Lumbala *et al;* 2013) In total, the CATT plate agglutination test and the SD Bioline HAT were performed on 88 samples. These two tests made it possible to detect specific antibodies LiTat 1.3 and LiTat 1.5 from *T. b. Gambiense* in all prospects.

The result was negative when a single-coloured band was observed on “C” in the result window. It was positive when two coloured bands were observed, either on line 1 and control line “C” (positive to Litat 1.3), or on line 2 and control line “C” (positive to Litat 1.5) or again when three coloured bands appeared in the control, 1 and 2 respectively, this means a positive result in Litat 1.3 and 1.5. The result was invalid when the control band “C” did not appear, regardless of the other results observed.

### Statistical analysis

Data were analysed using R.3.1.1 software. This analysis was used to calculate prevalence (p), sensitivity (Se) and specificity (Sp), positive predictive value (PPV) and negative predictive value (NPV). A P value of less than 0.05 was significant.

## Results

### General characteristics

This study involved 88 people recruited as follows in different foci, Campo 59.1% (n = 52), Yokadouma 31.8% (n= 28) and Fontem 9.1% (n = 8). The gender distribution in the foci investigated was 61.4% (n=54) male and 38.4% (n=34) female and the average age was 35.4 ± 19.0 years, ranging from 5 to 75 years.

### HAT seroprevalence

The serological prevalence (Figure 3) of HAT was 36.4% (95% CI: 27.1-46.7) with the whole blood CATT (32/88) and 18.2% (95% CI: 11.5-27.5) with the SD Bioline HAT (16/88). To increase the sensitivity of the CATT, successive dilutions of whole blood were performed. The threshold of suspicion of a case of HAT in Cameroon is represented by a positive CATT from 1:16 of diluted blood (PNLTHA, Cameroon) ^15^. Based on this national algorithm, our results suggested that the overall seroprevalence with the CATT method was 11.4% (95% CI: 6.3-9.7) in the probed foci (10/88).

**Figure 3.**
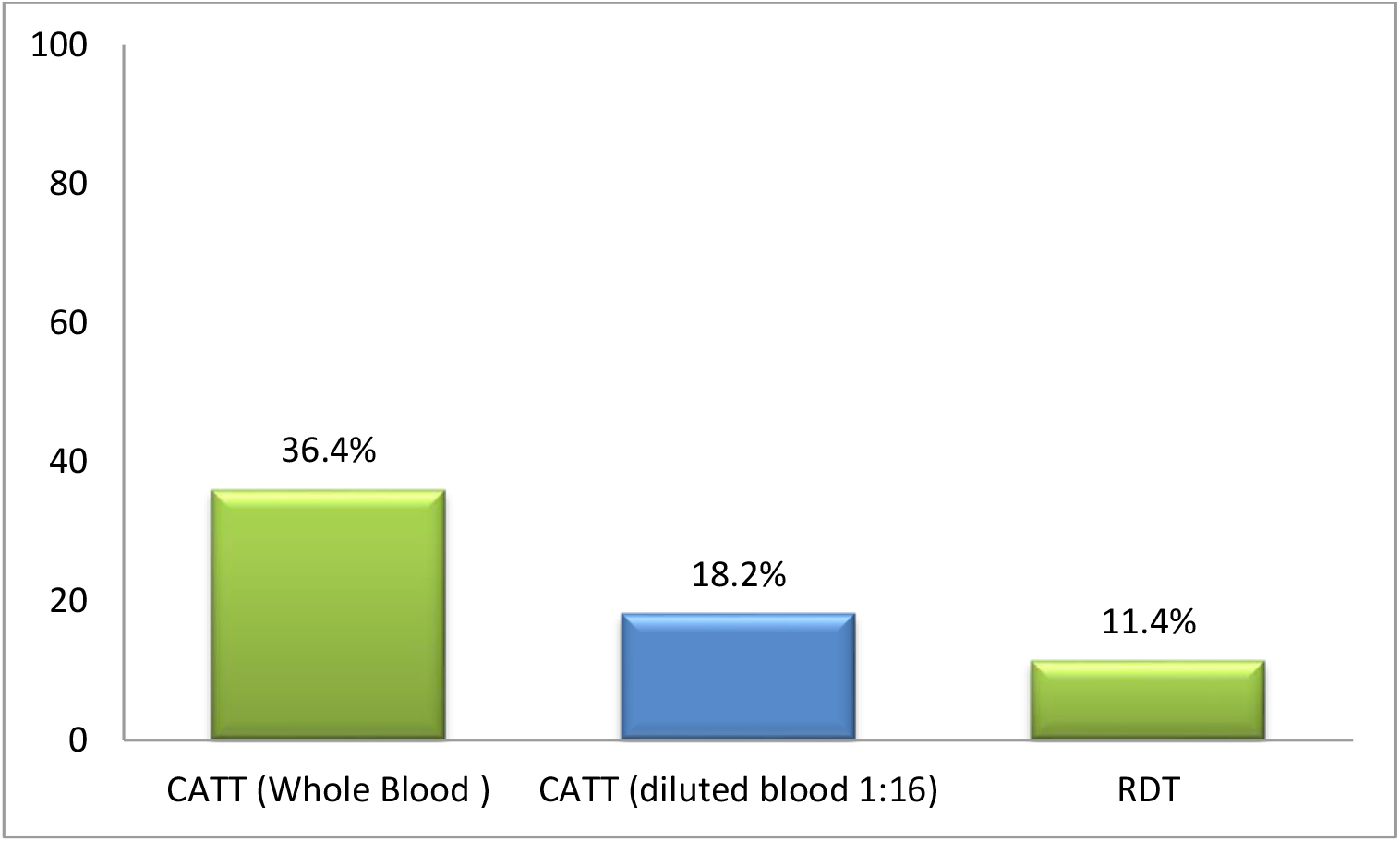
Serological prevalence. CATT ST: Card Agglutination Test for Trypanosomiasis with Whole Blood ; CATT 1:16: Card Agglutination Test for Trypanosomiasis diluted 1:16 ; RDT: Rapid Diagnostic Test

### Performance of rapid tests for HAT case detection

The performance of SD Bioline HAT has been calculated taking the CATT as the reference. We observe that, as the dilutions are made, the sensitivities of the RDT increase while the specificities decrease.

The threshold for CATT positivity for blood dilution is 1:16 in Cameroon. At this threshold, the SD Bioline HAT showed a sensitivity of 80.0%, a negative predictive value of 97.22% and the p-value of each was greater than 0.05 indicating a non-significant difference between these values and those of the CATT. At the same threshold, the specificity was 89.7% while the positive predictive value was 50.0% and the p-value of each was less than 0.05, showing a significant difference between the specificity of the CATT and the SD Bioline HAT is statistically. Table 1 shows the values of the intrinsic (Se and Sp) and extrinsic (PPV and NPV) performance indicators of SD Bioline HAT.

**Table 1.**
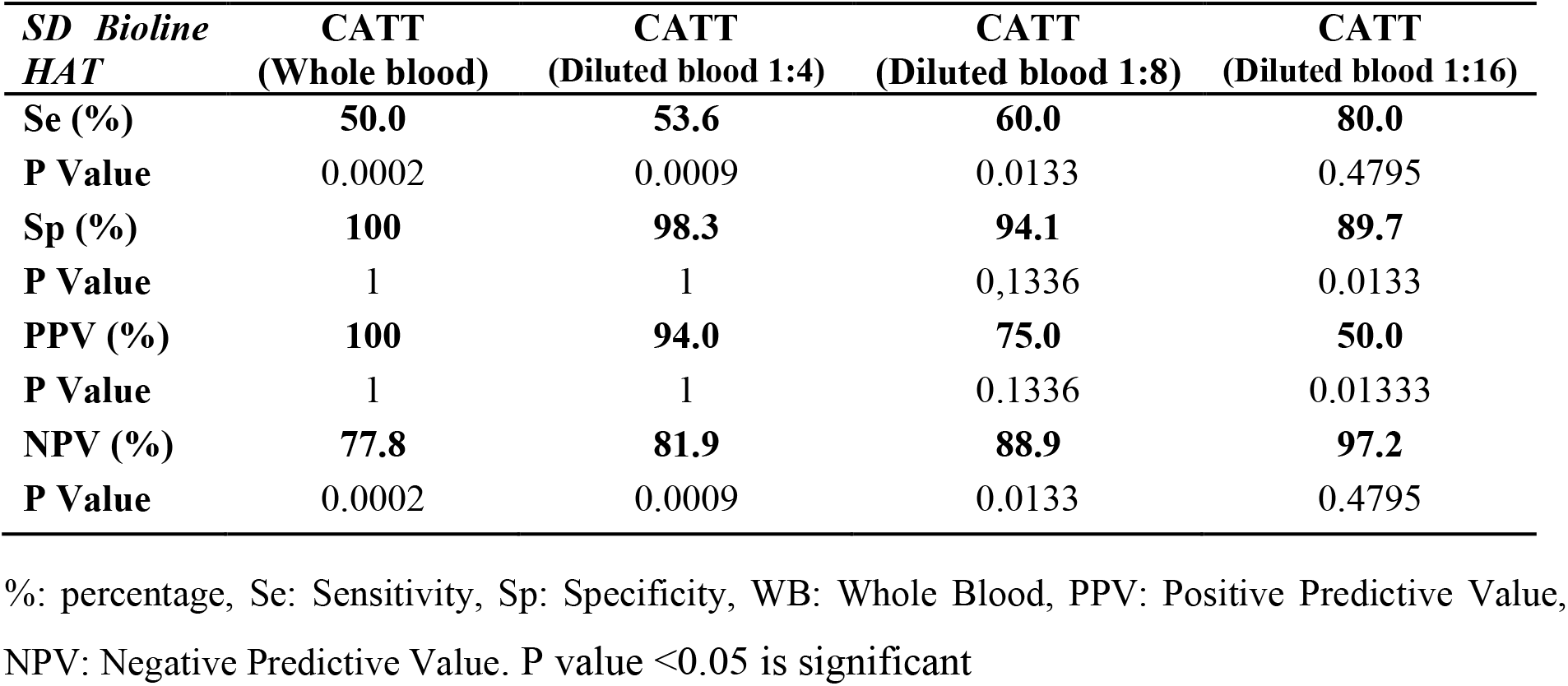
Descriptive statistics of the performance values of SD Bioline HAT.

| *SD Bioline HAT* | **CATT**<br>**(Whole blood)** | **CATT**<br>**(Diluted blood 1:4)** | **CATT**<br>**(Diluted blood 1:8)** | **CATT**<br>**(Diluted blood 1:16)** |
| --- | --- | --- | --- | --- |
| **Se (%)** | **50.0** | **53.6** | **60.0** | **80.0** |
| **P Value** | 0.0002 | 0.0009 | 0.0133 | 0.4795 |
| **Sp (%)** | **100** | **98.3** | **94.1** | **89.7** |
| **P Value** | 1 | 1 | 0,1336 | 0.0133 |
| **PPV (%)** | **100** | **94.0** | **75.0** | **50.0** |
| **P Value** | 1 | 1 | 0.1336 | 0.01333 |
| **NPV (%)** | **77.8** | **81.9** | **88.9** | **97.2** |
| **P Value** | 0.0002 | 0.0009 | 0.0133 | 0.4795 |

%, percentage, Se: Sensitivity, Sp: Specificity, WB: Whole Blood, PPV: Positive Predictive Value, NPV: Negative Predictive Value. P value <0.05 is significant

## Discussion

The main objective of our work was to evaluate the contribution of the rapid diagnostic test in the screening of HAT cases to *T. b. gambiense* in Cameroon. We had as a reference test the CATT where we performed four dilutions (1:2, 1:4, 1:8, 1:16) in addition to whole blood. We have used this RDTs because it is now available and distributed by the institute of tropical medicine for screening for HAT. To evaluate this test, samples were collected in three HAT outbreaks foci in Cameroon: Campo, Yokadouma and Fontem. Campo and Fontem are active and recognized HAT foci in Cameroon while the Yokadouma home is a suspect and silent focus ^16,17^. The investigation in the latter focus (Yokadouma) follows the movement of refugee victims of political instability in the Central African Republic (CAR), mostly coming from Nola, which is an active and recognized HAT focus in the CAR^18^.

Analyses with CATT included 88 individuals, the majority of whom were surveyed during Campo outbreak and the rest in the outbreaks of Yokadouma and Fontem.

The overall prevalence of serum HAT (positive CATT on whole blood) was 36%. It was reduced to 11.4% by considering the dilution of whole blood to 1:16 which corresponds to Cameroon’s threshold of suspicion of a case of HAT requiring surveillance ^15^. It should be noted that for HAT screening, WHO recommends that for any positive whole blood CATT, successive dilutions should be made in order to eliminate possible false positives.

In Cameroon, anyone with a diluted blood CATT ≥ 1:16 is considered to be serological HAT-positive and is being monitored by PNLTHA. This threshold of suspicion fixed to eliminate false positives varies from country to country and the epidemiological situation of HAT, Indeed, it is set at 1:8 in Chad and in the CAR, and 1:4 in the DRC ^15^. The false positivity of the CATT test may be due to cross-reactions following exposure of the individual to other pathogens, including animal trypanosomes: *T. b. Brucei, T. Congolense* ^19^. It has been shown that diseases such as schistosomiasis, filariasis or toxoplasmosis would be able to agglutinate CATT at low titres ^20,21^. Similarly, we observed the presence of microfilariae in the ganglion fluid of a CATT positive whole blood sample during our fieldwork.

Using SD Bioline HAT as an alternative screening tool, the total blood test positivity rate was 18.2%. However, at the blood threshold diluted 1:16, we obtained a prevalence of 11.4% closer to 18.2% found for the SD Bioline HAT test than undiluted blood. These values therefore indicate an approximation between the detection capability of CATT blood 1:16 and that of SD Bioline HAT.

The sensitivity of SD Bioline HAT is its ability to give positive results in all people who have been in contact with *T.b.gambiense* and have antibodies against Litat 1.3 and Litat 1.5 in their blood. The specificity in terms of its capacity has given negative results in all those who have never been in contact with *T.b.gambiense*. These two indicator values of the performance of the RDTs, to identify the serological cases of HAT were estimated by taking the CATT as a reference method.

The sensitivity of this RDT increased as CATT was diluted, unlike specificity. At the CATT blood threshold 1:16, the Se was 80.0% while Sp was 89.7%. These values suggest a satisfactory performance of SD Bioline HAT in the investigation of suspected cases of HAT. These results show some differences with the values found by Sternberg et *al*. (2014) the Se = 82%, Sp = 97%; by evaluating the performance of SD Bioline HAT and two prototype RDTs, out of 500 samples including 250 cases and 250 controls in Angola, CAR, and Uganda^22^. In addition, Bisser et *al*. obtained a sensitivity of 87.8% in the evaluation of the optimization of this same test with 49 confirmed parasitological THA specimens and 93-95% specificities following the evaluation of SD Bioline HAT and the SD Bioline optimized on 399 control samples in active screening in the DRC ^23^. They also obtained a sensitivity of 89.3% by comparing this RDTs with CATT diluted 1/8. The sensitivity of SD Bioline HAT which is 80% observed in this study was slightly low compared to that mentioned by the manufacturer (98%). The differences could be explained by our small sample size compared to theirs. In addition, we did not confirm parasitological cases.

### Limitations of the study

This study focuses on HAT disease, which is a rare and neglected tropical disease in Cameroon. The main limitations of this study were limited access to different collection foci, located in remote areas, small sample size, as well as financial limitations for the acquisition of rapid diagnostic tests.

## Conclusion

The study in three HAT foci in Cameroon revealed 18.2% of serological cases with the CATT method and 11.4% with SD Bioline HAT. The performance of SD Bioline HAT RDT compared to the CATT method shows that SD Bioline HAT could be an alternative to be adopted in most HAT foci in Cameroon. Considering that these foci are in remote areas, without appropriate infrastructure and technical platforms for perform CATT.

## Data Availability

All data produced in the present work are contained in the manuscript

## Acknowledgements

The study received support from the WHO office through grants to the national program for the control of human trypanosomiasis in Cameroon and from OCEAC for the acquisition and execution of rapid diagnostic tests for HAT. The authors present their tributes to the memory of Mr. Ebo’o Eyenga, coordinator of the National Program for Control of Human African Trypanosomiasis (PNLTHA) of Cameroon, who participated in the conceptualization and planning of this study in the field. Mr. Ebo’o died on October 13, 2018, during the drafting process of this paper.

## Notes

**Conflict of interest:** the authors declare no potential conflict of interest.

**Funding:** This study has not received any funding or external support

### Competing Interest Statement

The authors have declared no competing interest.

### Funding Statement

This study did not receive any funding

### Author Declarations

The ethical committee of the School of Health Sciences; Catholic University Of Central Africa gave ethical approval No 2015 / 0003 for this work.

### Summary of Updates

all sections have been revised

